# Implementing a Personalised Nutrition Service in a Real-Life Military Setting: Feasibility and Consumer Acceptance

**DOI:** 10.64898/2026.07.09.26357650

**Authors:** Sandra van der Haar, Meeke Ummels, Suzan Wopereis, Femke Hoevenaars, Imre W.K. Kouw, Martijn Noort

## Abstract

**Background:** Despite growing evidence that personalised nutrition (PN) can improve dietary behaviour and health outcomes, the implementation of PN food services in real-world settings remains largely unexplored. This study evaluated the feasibility of a comprehensive PN service for delivering personalised nutrition products (PNPs) in a military setting and assessed consumer acceptance following repeated consumption.

**Methods:** A complete PN service was developed to design, produce, and distribute personalised nutrition products (PNPs). Twenty-five participants from the Royal Netherlands Army took part in a feasibility study, using a single-cohort pre-test post-test design. Each participant consumed a PNP on 10 consecutive days during a two-week combat training exercise. Product liking was assessed through daily questionnaires, while broader user experiences were assessed at baseline (t=0), one week follow-up (t=1) and two-week endline (t=2).

**Results:** The PN service was succesfully implemented during a real-life military training exercise. In total, 229 PNPs were produced and distributed. Participants evaluated all aspects of the PN service from just below neutral to slightly positive (3.4 ± 1.6 to 5.0 ± 1.7 on 7-point scales). Across the different product combinations, mean overall liking and all sensory attributes were slightly positive (5.7 ± 1.7 to 6.1 ± 1.6 on 9-point scales). Overall liking did not change over time (*F* (9,194) = 1.242, *p*=0.27). Attitudes toward a PNP service in the army decreased over time, but remained just-above neutral at study end.

**Conclusions:** This study demonstrates the feasibility of a personalised nutrition service in a military setting. Both the service and PNPs received modestly positive evaluations, while repeated exposure did not change overall liking of the products. Although further optimisation of the service is warranted, these findings support the potential for broader implementation of PN services in military and other high-performance environments.

## 1. Introduction

Personalised Nutrition (PN) is founded in evidence-based science, and uses individual-specific data to improve dietary behaviours and health outcomes (Gevaert et al., 2020). Several studies have shown that PN interventions are more effective than generic interventions using a ‘one-size-fits-all’ approach in improving dietary habits (Carlos Celis-Morales et al., 2017; C. Celis-Morales et al., 2017; Jinnette et al., 2021; Rijnaarts et al., 2021) and health outcomes such as cardio-metabolic profile (Bermingham et al., 2024; Trouwborst et al., 2023; van der Haar et al., 2021) postprandial blood glucose levels (Zeevi et al., 2015) and gastro-intestinal complaints (Rijnaarts et al., 2022). A key challenge in translating PN into practice, is that most existing services are limited to providing dietary advice, meal plans, or grocery recommendations (Zhang et al., 2022). Emerging evidence suggests that embedding personalised recommendations directly into foods and food services may facilitate adherence and potentially enhance health impact compared to advice-only approaches (de Jong et al., 2026). However, the translation of individual health-related data into tailored food products that can be produced and delivered at scale remains largely unexplored and few studies have investigated how personalised nutrition products can be developed, delivered, and consumed within real-world environments. Within the IMAGINE research consortium (website IMAGINE - Health-Holland), this gap has been addressed by the development of a complete PN service that translates individual nutritional requirements into personalised in-between meals (Noort, M. *et al*. (2026) *manuscript in preparation*). The PN service integrates the generation of personalised dietary advice with the design, production, and distribution of customised in-between meals, thereby providing a unique opportunity to evaluate how PN can be implemented in practice (Noort et al., 2026). An important element of the service is three-dimensional (3D) food printing. As an additive manufacturing process, 3D food printing enables foods to be produced layer-by-layer according to a digital design, allowing nutrient composition, portion size, shape, and functionality to be tailored to individual requirements (Derossi et al., 2020; Sun et al., 2015; Zhang et al., 2022).

The military represents a particularly relevant setting in which to evaluate such an approach, as personnel often operate under physically and mentally demanding conditions that result in substantial variability in nutritional requirements (Tharion et al., 2005). Tailored nutrient compositions and functional ingredients may offer specific benefits for these specialised requirements. For example, physical and mental health issues are common in military personnel, such as gastro-intestinal problems (Wang et al., 2015), musculoskeletal injuries (Hauret et al., 2010) post-traumatic stress disorder, sleep problems and depression (Chou et al., 2016; Thomas et al., 2010). Providing optimal nutrition to soldiers through PN might therefore contribute to physical and mental health, including aspects such as improving vigilance and physiological recovery (Karl et al., 2022).

Beyond the technical feasibility, successful implementation of personalised nutrition services depends largely on consumers acceptance and sustained use. Previous research has shown that innovations in food have high failure rates (∼50%) (Michaut, 2004), partly due to unforeseen consumer reactions (Ronteltap & van Trijp, 2007). To be accepted, a new technology or novel food must demonstrate value and reassure consumers (Lupton 2017). For personalised nutrition specifically, acceptance is likely influenced by the perceived relevance and value of the tailored product to the individual user (Ross et al., 2022). Caulier was the first to investigate the perception of actual 3D-printed food products in a military setting. Their findings demonstrated that repeated consumption of a 3D-printed cereal-based recovery bar increased participant satisfaction and acceptance, and suggested that the perceived degree of personalisation contributed to these outcomes (Caulier et al., 2020). However, user evaluations of an integrated PN service encompassing product development, production, delivery, and consumption has not yet been performed.

Therefore, this study aimed to assess the feasibility of implementing a 3D food printing-based PN service in a real-life military environment. As consumer acceptance is an important determinant of successful implementation, product liking, consumer intentions, and attitudes toward personalised nutrition products (PNPs) following repeated consumption were assessed.

## 2. Materials and methods

### 2.1 Participants

Participants were recruited among different military units of the Royal Netherlands Army. In coordination with the army commanders, an infantry training unit of 40 soldiers that took place in and around a military base in Oirschot, the Netherlands was selected based on the criterium of being based at the same place for a duration of two weeks. Soldiers within this unit were approached by the researchers to participate in a field lab. An information meeting was organised before the start of the study, in which participants were informed, provided written informed consent and were checked for eligibility. Inclusion criteria were: Dutch speaking and _≥_16 years of age. Exclusion criteria included having food allergies or intolerances or other medical contra-indications as decided by a military physician.

A cohort of 25 soldiers enrolled for participation in a two-week study period, during which they were taking part in an infantry training. The training program prepared them for placement at a combat unit and consisted of offensive and defensive combat training. The first part of the training program (in week one of the study) took place at the military base of Oirschot, Harskamp and Stroe, with activities including practice of shooting at the shooting range, performing sports, and realistic combat training. The second part of the training program involved field training that took place in the woods close to the military base of Oirschot, where the soldiers were sleeping outside for three consecutive nights, with a maximum of four hours of sleep per night.

### 2.2 Study design

The study followed a single-cohort pre-test post-test design, with a duration of two weeks, and was executed between May 27^th^ and June 7^th^ 2024. In this proof-of-concept study, the PN service developed within the IMAGINE research consortium was evaluated in terms of consumer experience and product liking in a real-life setting. Next to a military setting, the service was also tested in a medical setting (Melchers et al., 2026 *manuscript under review).* The PN service contained several elements: 1) *a digital application* for consumers to provide personal data, indicate taste preferences, and to receive feedback on the PNPs they received 2) *a dietary advice algorithm*, in which the aforementioned personal data was used as input to generate dietary advice based on evidence-based knowledge rules 3) *a prototype Flexible Food Manufacturing Platform (FFMP) –* which is a mini food-factory using 3D food printing and dosing stations for various food materials and supplement powders - that fully automatically produced the PNP based on the dietary advice algorithm. The service as a whole produced fresh personalised in-between meals (Nutri-Bites) that were tailored to an individuals’ needs and preferences, containing nutrients that support evidence-based health benefits. The paper of Noort *et al* describes in detail how the PN advice system was designed and developed, including the knowledge base for the underlying personalised dietary advice, development of food materials for the PNPs and engineering of the prototype FFMP (Noort et al., 2026, *unpublished results*).

The intervention consisted of daily PNP consumption and concept and product evaluation using questionnaires for 10 consecutive working days (Figure 1). Before each working day, the personal participant data needed as input for the next day’s PNP was collected through the digital application. Weekend days were not included in the evaluation. As this was a feasibility study and the aim was to test the prototype and evaluate user-experiences in a real-life setting, no control group was included.

**Figure 1.**
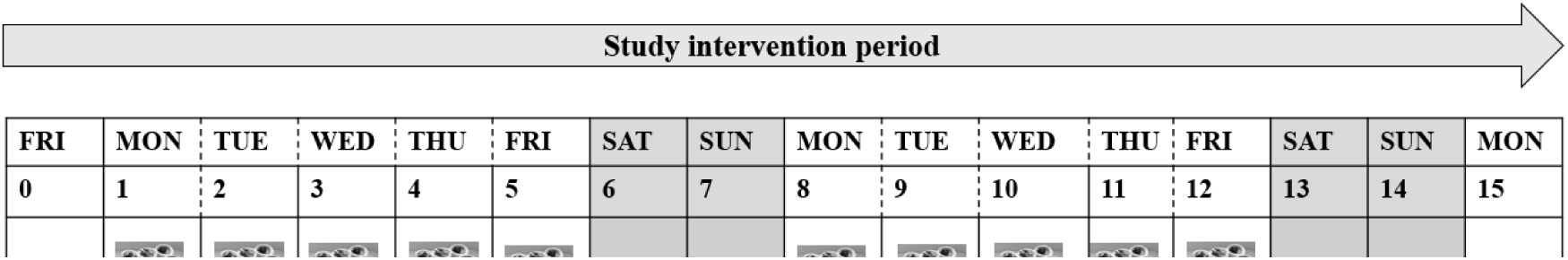
Overview of study design. In this 10 day study a PNP (personalised nutrition product) was distributed and assessed on weekdays in n= 25 soldiers by using daily questionnaires after product consumption. At baseline (t=0), after five days of PNP consumption(t=1) and at the end of the study period (t=2), a survey was administered to assess consumer attitudes, intentions, and perceptions toward personalised

### 2.3 Study procedures

During the study period, a FFMP was located at the TNO premises at the High Tech Campus in Eindhoven, the Netherlands, close to the military base. The FFMP produced daily PNPs (for participants referred to as “Nutri-Bite”) on the basis of a personalised dietary advice, summarised in a printer code. The advice underlying the PNP was generated with several fixed and dynamic input parameters. Body weight, height and wrist circumference were measured (for determining body type: ectomorph, mesomorph or endomorph) and sex, age, average daily physical activity level, vitamin D level, and habitual fiber intake were collected through a survey during a screening visit before study commencement as fixed parameters per subject. Dynamic parameters, such as specific daily Physical Activity Level (PAL) data, gastrointestinal complaints (GI complaints), coffee intake, sleep quality and taste preference were collected daily through a digital application (TNO Diamonds dashboard) that was accessible for participants on study tablets. Daily PAL-data was filled out in a physical activity planner by the group commander for each individual soldier and then transferred to the application. All the aforementioned parameters were used as input for the PNP on the following day. Total daily PAL was determined by combining general PAL with the individual daily PAL values for specific training activities over a 24-hour period, as recorded in a physical activity log provided by the group commander.

Each day, after all input parameters were collected, 25 printer codes were generated and sent to the FFMP. The PNPs were produced on the next morning between 08:00 AM and 1:00 PM. After production, the PNPs were transported to the military base in a cooled transportation box. The PNPs were distributed in disposable containers as an in-between meal product between lunch and dinner, and consumption always took place in the afternoon between 2:00 PM and 4:00 PM. Participants were informed about the research aims, but not on the exact production process of the PNPs that included the 3D-printer (incomplete disclosure) to minimise the risk of bias in the sensory evaluations. During product consumption, participants filled out a short questionnaire evaluating their PNP on several sensory aspects such as overall liking, taste, and texture. Furthermore, participants indicated how much of the product they had eaten (they were not obliged to consume the whole product) and reported GI complaints if any had followed after consumption of the PNP the day before. Besides these daily questionnaires, participants were asked to fill out a more detailed questionnaire at baseline (t=0), after five days of PNP consumption (t=1), and at the end of the study (t=2), assessing their attitudes, intentions, perceived benefits, and barriers towards a personalised food service in the army.

### 2.4 Personalised Nutrition Products (PNPs)

#### 2.4.1 Health benefits and nutritional advice

Baseline data profile, daily questionnaires, and daily physical activity level were integrated into selection of a ‘health benefit’ which served as input for the PNP of the next day. The pre-defined health benefits comprised of 1) immune health, 2) digestive health, 3) endurance, 4) muscle power, 5) recovery, and 6) mental performance. The daily health benefit selection protocol worked as follows: On the first day, participants were assigned based on either a high score (4-5 points) on the immunity questionnaire or a low fibre intake (less than 22 g per day according to the fibre screening questionnaire). If neither of these conditions applied, the health benefit that best matched the main activity planned for the day was chosen. For all consecutive days, assignment of the same health benefit was restricted to a maximum of two consecutive days; ‘recovery’ was mandatorily assigned on a day succeeding ‘muscle power’, and any extreme outcomes from daily questionnaires superseded activity-based selection criteria. Nutritional recommendations associated to each health benefits are described in **Supplemental Table 1a**. Micronutrients were selected based upon literature review taking the principles of the evidence pyramid as a guideline (Yetley et al., 2017). Nutrients were selected in case of presence of a health claim, meta-analysis or convincing evidence from randomised controlled trials. Macronutrient composition and sizing of the product was based upon the level of activity in case of muscle power, endurance or recovery. In cases digestion, immunity, or mental performance were assigned as health benefit, body type determined macronutrient composition and product size.

Participants received daily feedback in the online portal regarding the health benefit that was assigned for them for the next day. The information was presented to them as follows: “Tomorrow, you will receive a PNP that supports your immune system, as you mentioned you are feeling unwell *(example)”*

#### 2.4.2 Product design

Each PNP consisted of 1) *a casing*: an edible multi-compartment container with fillable cavities, made out of a variety of doughs or pastes 2) *a filling* which was fruit-, yoghurt-, custard-, cheese-, peanut-, or chocolate-based and 3) *a topping,* containing a flavor that was selected by the participant (chocolate, caramel, summer fruit, strawberry, green apple, orange, or lemon). In addition, micronutrient powders were dosed in the bottom of the casing cavities of the PNP, to fulfil a specific micronutrient composition. **Figure 2** shows a visual example of a PNP as presented to the participants. The product composition of each PNP (casing, filling, topping) was based on the macro- and micronutrient composition for each health profile as described in **Supplemental** Error! Reference source not found.**b**, which was in turn determined by the fixed and flexible input parameters specifically related to the individual participant and their specific activity and conditions. Some input parameters (such as body type, PAL data and dosing of a certain ingredient) also affected the size of the PNP (six cavities or eight cavities).

**Figure 2.**
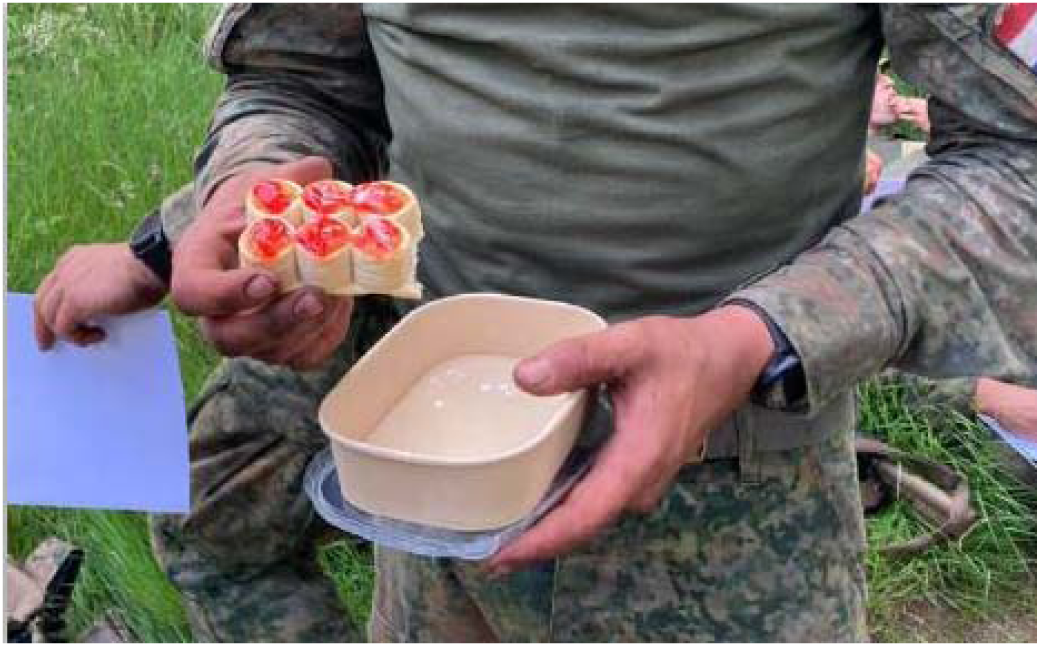
Photo of a PNP as presented to the participants, consisting of a baked dough casing, filling with incorporated micronutrients powder, and topping to match a specific macro- and micronutrient composition per health profile.

#### 2.4.2 Production process

PNPs were produced using the FFMP, a fully automated mini food-factory. Dough casings were 3D-printed and baked at 150 °C for 2, 4 or 12 minutes, according to dough type, then cooled to a core temperature below 40 °C. Micronutrient powders were dosed to the casing cavities, followed by the appropriate filling and participant selected topping.

Finished PNP’s were packed in a disposable carton box, labelled with a participant ID and stored at 5 °C, before transport in cooled boxes to the military base. All products were consumed within eight hours after production to ensure a fresh product texture and microbial safe product.

### 2.5 Study measures

#### 2.5.1 Product consumption and sensory evaluation

Several sensory parameters were daily assessed using a questionnaire evaluating the PNPs. Participants rated the PNPs on appearance, mouthfeel, taste, aftertaste, freshness, and overall liking on a nine-point Hedonic scale ranging from ‘dislike extremely (1)’ to ‘like extremely (9)’. After rating these attributes, participants filled out an open question (optional) on how the PNP could be improved in their opinion. Hereafter, participants were asked on product consumption (*I have consumed the product completely, ‘yes’ or ‘no’*) thereby indicating a reason in case of no complete consumption. Finally, participants were asked to indicate how likely it was they would be willing to consume this specific PNP again, on a seven-point Likert scale ranging from ‘very unlikely (1)’ to ‘very likely (7)’ and to fill out a reason (optional).

#### 2.5.2 Gastro-intestinal complaints

GI complaints following consumption of the PNP on the previous day were assessed daily, except for day one of the study, since the participants had not yet consumed a PNP. Participants were asked the following question: “*After consumption of the PNP yesterday, did you experience any gastro-intestinal complaints such as bloating, belching, nausea, flatulence, diarrhea or constipation?*” If they indicated yes, they were asked to describe which type of complaint they experienced.

#### 2.5.3 Attitudes, intentions, and perceptions towards personalised food service

At three time points, t=0, t=1, and t=2, participants completed questionnaires assessing their attitudes, intentions, and perceptions toward a personalised food service. All items were rated on a seven-point Likert scale. In the baseline questionnaire, the concept of the personalised food service was first explained to them as they had not yet seen it. Participants then completed items measuring their general attitude and perceived value towards a personalised food service in the army (1=not at all valuable, 7=very valuable), as well as their intention to adopt. Intention items reflected the three stages of behaviour change: pre-contemplation, contemplation, and action (Prochaska & DiClemente, 1982), assessed by using the following phrase: *I would consider to make use of it, I plan to make use of it, I certainly would make use of it* (1=completely disagree, 7=completely agree).

At both t=1 and t=2 questionnaires, the same attitude and intention items were repeated to assess changes over time. In addition, participants rated their expectations regarding potential health benefits on PNP consumption (e.g., on digestion, alertness, immune system, recovery, endurance, and muscle strength). Finally, they also evaluated potential benefits and barriers of PNPs, including aspects such as taste, freshness, feasibility of implementation within the army setting, time investment, and use of personal data.

#### 2.5.4 Self-rated health and wellbeing

At t=0, t=1, and t=2 participants were asked to rate their physical and mental health. Self-perceived overall health was measured with the Self-Rated Health scale, a widely used single-item measure developed by Ware & Sherbourne (1992), and encompassed the following question: “*How would you currently rate your own health on a scale from 0 to 100, where 0 is very poor health and 100 is excellent health*”? Mental health (wellbeing) was measured with the World Health Organization (WHO)-5 Well-being Index. This index consists of five statements on wellbeing referring to the past two weeks. Each statement is rated on a 6-point scale, ranging from ‘at no time (0)’ to ‘all of the time’ (5) with higher scores indicating better wellbeing. The scores on each statement make up a sub score, that is multiplied by 4 to give a total score, with 0 representing the worst imaginable wellbeing an 100 representing the best imaginable wellbeing. Total scores below 50 indicate poor wellbeing.

#### 2.5.5 Demographics and other study sample characteristics

From the baseline questionnaire that was used as input for the PNP, socio-demographic characteristics of the participants were collected such as sex, age, body mass index (BMI), and habitual fibre intake. Furthermore, habitual in-between meal behaviour (frequency and type of food products) and food choices motives of all participants were included from the t=0 questionnaire. Consumer choice was assessed from a pre-defined list of 12 motives (healthy, animal-friendly, safe, natural, easy to use, affordable, ethical concern, sensory appealing, familiar, mood, environment, and weight management) where participants were asked on their motive when buying food products on 7-point Likert scales, ranging from ‘very unimportant (1)’ to ‘very important (7)’ (Steptoe, 1995).

Habitual in-between meal consumption was assessed with questions on the frequency of eating in-between meals (‘yes’ or ‘no’) and if yes, to indicate how many in-between meals they consumed on average per day.

### 2.6 Statistical analyses

Data on PNP composition and sensory ratings were presented as means ± SD. Categorical variables (GI-complaints, product consumption) were displayed as frequencies and percentages. A general linear model (GLM) with Time as fixed factor and Participant as random factor was conducted to assess the effect of repeated consumption on overall liking scores of all PNPs. Since this was a feasibility study with limited numbers of observations for each combination of product characteristics, the assumptions to test the effect of repeated consumption of specific filling, casing, and topping combinations in a linear mixed model were not met. To test the difference in mean Likert-scores over time (t=0, t=1, t=2) for attitudes, intentions, self-rated health and wellbeing, repeated measures ANOVA was applied. Subsequently, Least Significant Difference (LSD) post-hoc tests were performed to assess the differences between timepoints. To test the difference in mean Likert-scores for benefits and barriers of PNPs between t=0 and t=2, paired samples t-tests were applied.

Sensory ratings of drop-outs were included in the sensory analyses and in the GLM. In the over-time analyses of the t=0, t=1, and t=2 questionnaires, drop-outs were excluded. Statistical significance was set at *p*<.05 for all analyses. All statistical analyses were performed using SPSS software (version 25.0; IBM Corp).

## 3. Results

### 3.1 Participants and baseline characteristics

A total of 25 male participants were included in the study with a mean age of 20.7±2.0 y [18-25] and BMI 24.1±1.9 kg/m^2^ [21.2-27.6]. Habitual fibre intake at baseline averaged 14.8±7 g/day, which is below the Dutch recommendations for males i.e. 40 g/day. The majority of participants (92%) was used to eating in-between meals in their daily diet with an average of 2.7±1.3 in-between meals per day. Based on the pre-defined list of 12 Food Choice Motives, the top five most important motives for food choice were: sensory appealing (6.0± 1.1), ‘gives me a good feeling’ (5.7±1.1), affordability (5.5±1.4), safety (5.3±1.7), and healthiness (5.3± 1.0).

Five participants (20%) dropped out during the two-week study period, which coincided with the most intensive sleep deprivation phase of the military training. All drop-outs reported not liking the study products as main reason for quitting the study, in combination with experiencing GI complaints (n=2/5) or having a high expectation of personal health benefits that were not met (n=1/5).

### 3.2 Implementation of the PN service

#### 3.2.1 Product consumption

The PN service was succesfully implemented during a military combat training exercise of two weeks, and a total number of 229 PNPs were produced and distributed to participants during the study on 10 subsequent working days. Of these, 208 PNPs (91%) were consumed entirely and 21 PNPs (8%) were consumed partly.

#### 3.2.2 Evaluation of the PN service

At the end of the two-week period, participants scored just above neutral for the number of choices in topping flavours (4.5±1.4), the time between ordering and distribution of the PNP (4.6±1.2), and the amount of personal information that had to be shared to produce the PNP (4.8±1.5). They were slightly positive on the amount of time spent on ordering a PNP (5.0±1.7) and on the user-friendliness of the application for ordering (5.0±1.7). Only for variation in PNPs, participants scored below the neutral score (3.4±1.6).

### 3.3 Product evaluations

**Table 1** shows the sensory evaluation of all distributed PNPs, without making a distinction in product characteristics. The mean scores for overall liking, appearance, mouthfeel, taste, aftertaste, and freshness were all slightly positive (ranging from 5.7±1.7 to 6.1±1.6). Willingness to consume the specific PNP again, received a neutral score (4.2±1.7).

**Table 1.** Sensory evaluation on taste and texture attributes, and overall liking for all PNPs (n=229).

| Sensory aspect | Mean score (SD)<sup>1</sup> |
| --- | --- |
| Overall liking | 5.9 (1.6) |
| Appearance | 6.0 (1.3) |
| Mouthfeel | 5.8 (1.6) |
| Taste | 6.1 (1.6) |
| Aftertaste | 5.7 (1.7) |
| Freshness | 6.0 (1.4) |
| Willingness to eat again | 4.2 (1.7)<sup>2</sup> |

<sup>1</sup>Assessed on a 9-point Hedonic scale, ranging from extremely unappealing (1) to extremely appealing (9).
<sup>2</sup> Assessed on a 7-point Likert Scale, ranging from not at all (1) to very much (7).

**Table 2** displays the overall liking score per specific casing and filling combination. Overall liking scores of the PNPs for specific casing and filling combinations ranged from 2 to 9. The protein casing combined with custard filling received the highest rating (8.4±0.5), while the plum paste combined with a peanut butter filling received the lowest rating (5.3±1.4).

**Table 2.** Overall liking score for specific casing and filling combinations within PNPs.

| Casing type<br>(dough/paste) | Filling type | Observations (n) | Mean score (SD)<sup>1</sup> |
| --- | --- | --- | --- |
| Protein | Custard | 8 | 8.4 (0.5) |
| Protein | Peanut butter | 51 | 5.7 (1.5) |
| Carrot | Ganache | 24 | 5.5 (1.7) |
| Plum | Peanut butter | 32 | 5.3 (1.4) |
| Plum | Custard | 21 | 5.7 (1.6) |
| Cereal | Yoghurt | 15 | 5.6 (1.4) |
| Cereal almond | Apple | 26 | 5.9 (1.8) |
| Cereal almond | Custard | 4 | 7.0 (0.0) |
| Cereal high fiber | Peanut butter | 26 | 6.0 (1.5) |
| Cereal high fiber | Banana-strawberry | 15 | 7.2 (1.2) |

<sup>1</sup>Assessed on a 9-point Hedonic scale, ranging from extremely unappealing (1) to extremely appealing (9). The addition of powders containing specific micronutrient combinations was not taken into account

#### 3.3.1 Overall liking over time

The overall liking scores per day were reported within a range of 5.4 ±1.5 (lowest mean score on day 1) and 6.5±1.8 (highest mean score on day 9) on a 9-point Hedonic scale (**Figure 3**). However, no time effect on overall liking scores was detected (*F* (9.194) = 1.242, *p* = 0.27).

**Figure 3.**
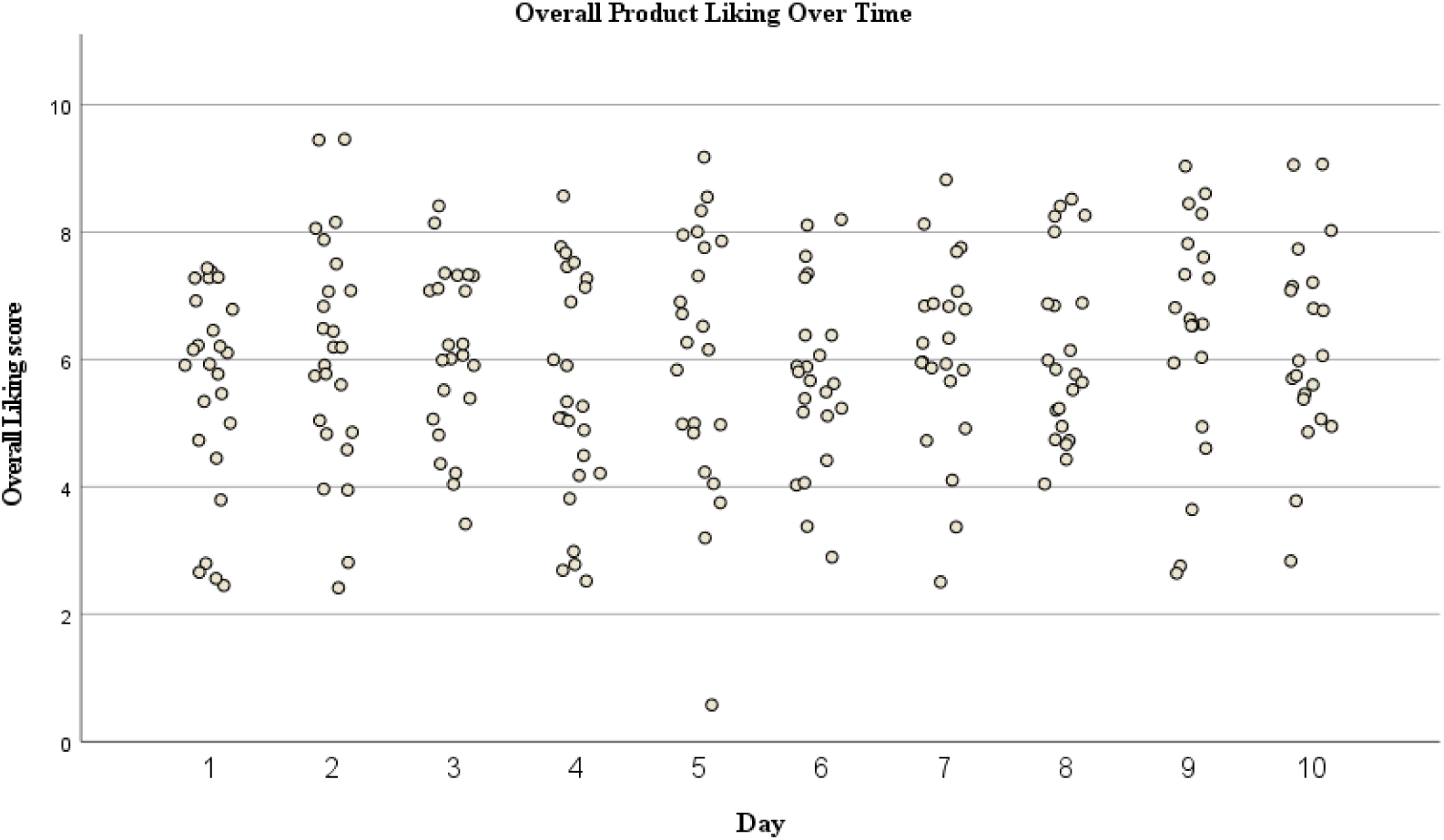
Overall product liking assessed daily after PNP consumption

#### 3.3.2 Attitudes, intentions, and perceptions during the intervention period

**Table 3** shows the mean scores per time point for consumer attitudes, intentions, and perceived benefits and barriers. The mean score for attitude towards PNPs in the army significantly decreased over time (*F* (2.38) = 11.5, *p*<0.001), even though mean scores were still above the neutral score at the end of the intervention period. Intentions to adopt PNPs decreased significantly over time, for the pre-contemplation stage (*F* (2,34) = 13.2, *p* < 0.001), contemplation stage (*F* (2,34) = 5.1, *p* = 0.01), and action stage (*F* (2,34) = 8.8, *p* < 0.001).

**Table 3.** Consumer attitudes, intentions to adopt, and perceived benefits and barriers, measured at baseline, after one week, and after two weeks.

| Survey item | Mean score (SD) on 7-point Likert Scales |  |  |  |
| --- | --- | --- | --- | --- |
|  | t=0 weeks | t=1 week | t=2 weeks | P value |
| *Attitude towards PNPs* |  |  |  |  |
| ‘A personalised food service in the army is valuable’ | 5.9 (1.0)<sup>a</sup> | 5.0 (1.4)<sup>b</sup> | 4.5 (1.6)<sup>b</sup> | **<0.001** |
| *Intention to adopt PNPs* |  |  |  |  |
| ‘I would consider it’ (pre-contemplation stage) | 5.9 (1.1)<sup>a</sup> | 5.4 (1.1)<sup>b</sup> | 4.3 (1.9)<sup>c</sup> | **<0.001** |
| ‘I am planning to’ (contemplation stage) | 5.7 (1.1)<sup>a</sup> | 4.9 (1.2)<sup>b</sup> | 4.6 (1.9)<sup>c</sup> | **0.01** |
| 'I will definitely' (action stage) | 5.8 (1.2) <sup>a</sup> | 4.8 (1.4) <sup>b</sup> | 4.0 (2.2) <sup>c</sup> | **<0.001** |
| *Perceived benefits and barriers of PNPs* |  |  |  |  |
| PNPs are tasty | 4.8 (1.2) |  | 4.4 (1.3) | 0.35 |
| PNPs are fresh | 5.3 (1.0) |  | 4.7 (1.1) | 0.06 |
| PNPs are difficult to implement in the army | 4.1 (1.5) |  | 5.0 (1.5) | 0.08 |
| PNPs are time-consuming | 3.4 (1.6) |  | 4.4 (1.1) | **0.02** |
| I have concerns about sharing my personal data for PNPs | 2.3 (1.4) |  | 2.4 (1.3) | 0.88 |

<sup>a,b,c</sup> Different letters indicate differences between time points, $P < 0.05$ .

Participants were on average slightly positive about the tastiness of the PNPs at baseline, and this did not differ after the two-week intervention period (*t=*0.95*, p* = 0.35). The same holds for freshness of the PNPs, although a trend for a lower score after the intervention was observed (*t =* 2.1*, p* = 0.06). At the start of the intervention, participants scored PNPs not to be time-consuming, however this increased over time to being more neutral (*t =* −2.6*, p* = 0.02). Participants were neutral on whether it would be difficult to implement PNPs in the army at the start of the intervention, and at the end of the intervention they slightly agreed to this statement (*t=-1.84, p=0.08).* Participants had no concerns on sharing their personal data to produce PNPs at the start of the study, and this did not change over time (*t =* −0.160*, p =* 0.88).

**Table 4** shows the mean scores per time point for attitudes toward potential health benefits of PNPs, self-rated health, and wellbeing. Mean scores for attitudes toward potential health benefits of PNP reduced significantly over time for PNPs with health profile immune health (*F* (2,36) = 3.7, *p* = 0.04), recovery (*F* (2,36) = 11.9, *p<*0.001), endurance (*F* (2,36) = 7.0, *p* = 0.009), and muscle power (*F* (2,36) = 15.5, *p<*0.001). For all of these health profiles, participants scored on average (slightly) positive at baseline, and more neutral toward the end of the study. For the health profile digestion, participants were on average slightly positive (5.0 ± 1.1) and this did not change over time (*p*=0.27). For the health profile alertness, the mean score was also slightly positive at baseline (4.7±1.5), and more neutral toward the end of the study (3.8±1.5), with a nearly-significant decrease over time (*p*=0.06).

**Table 4.** Attitudes toward potential health benefits of PNPs, self-rated health and wellbeing, measured at baseline, one week after the intervention, and two weeks after the intervention.

| Survey item | Mean scores (SD) |  |  | P value |
| --- | --- | --- | --- | --- |
|  | t=0 weeks | t=1 week | t=2 weeks |  |
| I think PNPs can have benefits for my... <sup>1</sup> |  |  |  |  |
| Digestive health | 5.0 (1.1) | 4.5 (1.3) | 4.8 (1.1) | 0.27 |
| Alertness | 4.7 (1.5) | 4.1 (1.6) | 3.8 (1.5) | 0.06 |
| Immune health | 5.1 (1.4) <sup>a</sup> | 4.2 (0.9) <sup>b</sup> | 4.2 (1.2) <sup>b</sup> | **0.04** |
| Recovery | 5.7 (1.2) <sup>a</sup> | 4.7 (1.1) <sup>b</sup> | 4.2 (1.1) <sup>b</sup> | **<.001** |
| Endurance | 5.2 (1.5) <sup>a</sup> | 3.9 (1.5) <sup>b</sup> | 4.1 (1.4) <sup>b</sup> | **0.009** |
| Muscle power | 5.3 (0.9) <sup>a</sup> | 3.9 (1.5) <sup>b</sup> | 4.1 (1.4) <sup>b</sup> | **<0.001** |
| Self-rated health score <sup>2</sup> | 78 (12) | 79 (11) | 76 (11) | 0.40 |
| Well-being index score <sup>3</sup> | 63 (11) <sup>a</sup> | 65 (15) <sup>a</sup> | 53 (10) <sup>b</sup> | **0.002** |

<sup>1</sup> Measured on 7-point Likert scales, ranging from strongly disagree (1) to strongly agree (7)
<sup>2</sup> Measured on a scale ranging from 0 (worst possible health) to 100 (best possible health)
<sup>3</sup> Index score ranging from 0 (worst possible quality of life) to 100 (best possible quality of life)
<sup>a,b</sup> Different letters indicate differences between time points, $P < 0.05$ .

Self-rated health scores remained stable over time, with mean values of 78±12 at baseline and 76±11 after two weeks of the intervention ((*F* (2,32) = 0.9, *p* = 0.40). In contrast, well-being index scores increased throughout the intervention period, rising from 53±10 to 63±11 (*F* (2,34) = 7.8, *p* = 0.002).

## 4. Discussion and conclusion

### 4.1 Principal findings and discussion

This study demonstrated the feasibility of implementing a 3D food printing-based personalised nutrition service in a real-life military setting. A complete service encompassing personalised dietary advice, product design, production, distribution, and consumption was successfully operationalised within a population characterised by substantial variation in nutritional requirements. In total, 229 personalised nutrition products were delivered during a demanding military training period, providing proof-of-concept that individual nutritional recommendations can be translated into tailored food products and implemented under real-world conditions. As consumer acceptance is a critical determinant of successful implementation, users’ experiences with both the products and the service were evaluated. Consumer acceptance of the personalised nutrition products was modestly positive. Overall liking and sensory attribute ratings were slightly above neutral for all product combinations, indicating that participants generally accepted the products despite their novelty and the demanding context in which they were consumed. Importantly, none of the evaluated product combinations received negative ratings. Due to the personalised nature of the intervention, participants received products that differed in characteristics, making direct sensory comparisons challenging. Nevertheless, grouping products according to casing and filling combinations showed differences in liking between product types, suggesting opportunities for further optimization of some product formulations.

A limited number of studies have previously conducted sensory evaluations of 3D-printed food products (Caulier et al., 2020; Khemacheevakul et al., 2021; Talens et al., 2022). In a consumer study among military personnel, Caulier et al. (2020) reported comparable overall liking scores for 3D-printed cereal-based recovery bars, ranging from 5.7 to 5.9 on 9-point hedonic scales, depending on bar type (Caulier et al., 2020). These scores are similar to our mean overall liking score of 5.9 for the PNPs in the present study. Talens and Rios (2022) evaluated 3D-printed breakfast bars in a panel of Spanish older adults. While concept and appearance were initially rated positively (6.0±2.0 on a 9-point scale), acceptability decreased significantly to a neutral score after tasting (5.0±2.0), suggesting that the sensory experience did not meet expectations of the product concept. Khemacheevakul et al. (2021) evaluated the sensory profile of several sugar-reduced and non-sugar-reduced 3D-printed chocolates. Liking scores ranged from 6.4±1.6 to 6.9±1.5. Their results showed that layering high- and low-sugar chocolate via 3D printing enabled up to 19% sugar reduction without adversely affecting perceived sweetness or overall liking. Considering that the PNPs in our study were unfamiliar products produced through an innovative service, and that consumers may initially approach such products with some degree of scepticism, the slightly positive sensory evaluations observed can be considered as a promising outcome at this stage (Popa & Popa, 2012).

Beyond product acceptance, participants provided generally favourable evaluations of implementation-related aspects of the PN service. Time investment, the amount of personal information that had to be shared, and the user-friendliness of the digital ordering application were evaluated around or above neutral. Previous research has shown that one of the challenges of PN services is that consumers may perceive the provisioning of personal information as time-consuming. In addition, user-friendliness has been identified as an important prerequisite for consumer engagement in personalised dietary advice tools and services (König et al., 2021; van der Haar et al., 2023). In the present study, participants did not perceive either the time investment, or the user-friendliness of the PN service as barriers, which is an important factor for implementation of this PN service. One implementation challenge identified in the present study was the limited perceived variation in products throughout the intervention period. This was primarily attributable to constraints of the prototype set-up, as no alternative casings and fillings with comparable nutritional properties had been developed, rather than to inherent limitations of the 3D printing technology itself. Consequently, day to day product variation depended largely on the training program and the practical feasibility of the pilot study. Since the training program mainly contained heavy physical exercise, the nutrient-dense peanut butter filling was advised most often. Future implementation efforts should therefore prioritize an increase in dietary variety while maintaining nutritional tailoring.

A relatively high prevalence of GI-complaints was reported following PNP consumption, affecting 36% of all participants. However, these findings should be interpreted with caution as total dietary intake of participants was not controlled during the study period and the PNP constituted only a small proportion of participants’ overall food intake. In addition, it is likely that factors other than PNP consumption contributed to GI-discomfort. The intervention period coincided with an intense training program, characterised by sleep deprivation and strenuous physical activities, both of which are known to affect gastrointestinal function. GI complaints are commonly reported among soldiers during military missions and training exercises (Wang et al., 2015). Finally, participants’ habitual low fibre intake at baseline may also have contributed to the occurrence of GI-discomfort during the intervention period. Nevertheless, monitoring tolerance and GI complaints remains important during future implementation studies.

Contrary to expectations, repeated consumption did not result in increased product liking over time. Although mean overall liking scores were descriptively higher at the end of the intervention than at baseline, no significant effect of time was observed. This finding differs from previous military research on 3D-printed foods (Caulier et al., 2020). However, unlike earlier studies in which participants repeatedly consumed the same product, the current intervention exposed participants to multiple personalised product variations. Consequently, familiarity with any single product remained limited, potentially attenuating a repeated-exposure effect.

Previous research on novel foods and food technologies, suggests that repeated exposure can increase familiarity, which in turn may positively affect consumers’ attitudes (Bruhn, 2007; Lyndhurst, 2009). In the present study, participants reported positive attitudes at baseline, however the mean scores for perceived added value and intentions to adopt across all three behavioural-stages (e.g. pre-contemplation, contemplation, action) decreased over time toward a more neutral or slightly positive score at the end of the study. A similar pattern was observed for perceived potential health benefits of both the products and the service. Ratings decreased from slightly positive at baseline, to more neutral at the end of the study for immune health, recovery, endurance, and muscle power. Only perceived digestive health benefits remained slightly positive throughout the intervention period. The observed may be explained by initially high expectations regarding personal health benefits, even though this was out of scope for this study. Importantly, participants remained open toward personalised nutrition, suggesting that real-life exposure did not undermine acceptance of the concept itself.

An encouraging finding for future implementation was the absence of substantial concerns regarding sharing personal information for the creation of personalised food products. Although privacy risks are frequently cited as a barrier to the adoption of PN, and previous studies have shown that many consumers perceive data sharing as a risk that outweighs potential benefits (Berezowska et al., 2015; Reinders et al., 2020) this was not observed in the present sample. Although this finding may partly reflect the military context and should be interpreted with caution, it suggests that data-sharing requirements are unlikely to constitute a major implementation barrier in similar settings.

Finally, interpretation of the findings should consider the challenging operational context in which the intervention was conducted. Participants underwent an intensive training programme, characterised by prolonged physical exertion, outdoor exercise and severe sleep restriction. These circumstances may have influenced both wellbeing and product evaluations. However, simultaneously the feasibility of deploying and evaluating a PN service under realistic military conditions has been demonstrated.

### 4.2 Strengths, limitations and recommendations for further research

A major strength of this study was the evaluation of a personalised nutrition (PN) service under real-world conditions in a military setting. Rather than assessing products in a controlled laboratory environment, participants interacted with the service and consumed the PNPs during their regular training activities. This approach provided insight into the feasibility of implementing a PN service in practice and generated user experiences that are likely to reflect real-world use and acceptance.

Another strength lies in the evaluation of the whole PN service. All components were tested and assessed, ranging from the personalised advice itself, ordering a PNP including a flavour selection, to the production of the PNPs and the subsequent sensory experience. Finally, the study design incorporated repeated exposure to PNPs, which provided deeper insights into longer-term user perceptions. Unlike single-exposure studies, repeated exposure allowed assessments of changes in product appreciation and attitudes toward the service over time.

Several limitations should also be acknowledged. First, this feasibility study evaluated an early prototype of the PN service, and further optimisation of both the products and service components is warranted. Product variety was limited, partly because food materials were developed to serve multiple target populations rather than being optimised specifically for military personnel (Noort et al., 2026, unpublished results). In addition, the FFMP was able to produce a limited number of 25 products per day, and therefore the service could only be evaluated in a relatively small sample. Following further optimisation of the platform, future studies should include larger study populations to improve statistical power and generalisability. Second, no control group was included. While this is common in early-stage feasibility research, the absence of a control condition limits conclusions regarding the added value of the PN service compared with conventional nutrition support. In future research, a semi-placebo design could be considered by comparing the PN service and personalised products with the provision of generic in-between meals and non-personalised nutritional advice. Third, user experiences were assessed using subjective questionnaires, which are inherently subject to limitations, such as response bias and socially desirable answering behaviour (Fisher, 1993; Podsakoff et al., 2003). Furthermore, the personalised nature of the intervention created challenges in the statistical analyses. PN is fundamentally a N-of-1 approach, whereas conventional statistical methods rely on mean differences within or between groups. Because product allocation depended on individual nutrition requirements, participants received different product combinations and differed in the frequency with which products were consumed. This unbalanced design limited the ability to examine changes in liking for individual product types over time. Another limitation concerns the incomplete disclosure of 3D-printing technology. Although extensive details regarding the production process were not provided during the intervention, all participants were aware that the products had been 3D-printed. This awareness may have influenced objective sensory evaluations. Previous research has shown that novel food technology neophobia can affect willingness to try novel foods and influence product evaluations (Caulier et al., 2020; Ross et al., 2022) However, military personnel have been reported to perceive 3D-printed foods relatively positively, and to be less susceptible to such neophobia (Caulier et al., 2020).

Finally, the present study focused on implementation feasibility and consumer acceptance, rather than efficacy. Controlled intervention trials are therefore required to assess the added value of PN for health improvement, particularly as consumers appear more receptive to 3D-printed foods and other food innovations when clear personal health benefits are evident (Silva et al., 2024). In a military context, it is important to study the effects of PN on both physical and cognitive performance, as these domains are central to the rationale of our PN products, and yet remain insufficiently supported by randomised clinical trials. Generating such evidence is essential to determine whether personalised approaches can significantly improve operational readiness and resilience in demanding military environments.

### 4.3 Conclusion

In conclusion, this study demonstrated the feasibility of implementing a full PN service in a real-life military setting. The successful integration of personalised dietary advice with the design, production, and distribution of tailored food products provides an important proof-of-concept for personalised nutrition in practice. Consumer acceptance was modestly positive, with PNPs receiving slightly favorable evaluations throughout the intervention. Although attitudes toward the service and its perceived benefits declined over time, they remained neutral to slightly positive at study end. Overall, these findings suggest that personalised nutrition services can be successfully delivered in demanding real-world environments. While further optimisation is needed, this study is an important step towards broader implementation. Future research should evaluate larger-scale deployment and assess the impact of such services on health and performance outcomes.

## Data Availability

All data produced in the present study are available upon reasonable request to the authors

## Declarations

### Ethics approval and consent to participate

All participants provided signed informed consent for inclusion in the study. Ethical clearance for the study was obtained from the WUR Research Ethics Committee (REC) under approval number 2024-041.

### Availability of data and materials

The datasets used and analysed during the current study are available from the corresponding author on reasonable request.

### Authors’ contributions

- Conceptualisation: SvdH, MU, FH, SW, MN
- Data curation: SvdH, MU
- Formal analysis: SvdH
- Investigation: SvdH, MU
- Methodology: SvdH, FH, MN, MU, SW, IWKK
- Writing – original draft: SvdH, MU, FH
- Writing – review and editing: IWKK, MN, SW

## Acknowledgements

The authors would like to express their sincere gratitude to Mark Tamis (Ministry of Defence) for his efforts in recruiting participants within the Royal Netherlands Army and for his support throughout the execution of the study. Furthermore, the authors would like to thank José van Uden (TNO), project manager of the IMAGINE consortium, for her coordination and supervision, Marc Hoppenbrouwers, André Rijfers, Roland Zegers, Dolf Klomp, Peter Giesen and Martyasa Putra for engineering and developing the FFMP, Eugene van Someren for implementing the digital ordering system for the PNP, Hannah Eggink and Lotte Peters for their support in developing and formulating the health nutritional profiles and Edwin Stam and Nerea Marina Larrañaga for producing the PNP’s (all TNO). Furthermore, we would like to thank Bei Tian, Seyed-Ali Ghoreishy and Jolanda Henket for developing and preparing the food materials. Lastly, we are grateful to Merel van Efferen for her contribution in the development of PNPs and Caya Lindner for her assistance in data collection and preparation (all WUR).

## References

Berezowska, A., Fischer, A. R. H., Ronteltap, A., van der Lans, I. A., & van Trijp, H. C. M. (2015). Consumer adoption of personalised nutrition services from the perspective of a risk–benefit trade-off. Genes & Nutrition, 10(6), 42. 10.1007/s12263-015-0478-y

Bermingham, K. M., Linenberg, I., Polidori, L., Asnicar, F., Arrè, A., Wolf, J., Badri, F., Bernard, H., Capdevila, J., Bulsiewicz, W. J., Gardner, C. D., Ordovas, J. M., Davies, R., Hadjigeorgiou, G., Hall, W. L., Delahanty, L. M., Valdes, A. M., Segata, N., Spector, T. D., & Berry, S. E. (2024). Effects of a personalized nutrition program on cardiometabolic health: a randomized controlled trial. Nat Med, 30(7), 1888–1897. 10.1038/s41591-024-02951-6

Bruhn, C. M. (2007). Enhancing consumer acceptance of new processing technologies. Innovative Food Science & Emerging Technologies, 8(4), 555–558.

Caulier, S., Doets, E., & Noort, M. (2020). An exploratory consumer study of 3D printed food perception in a real-life military setting. Food Quality and Preference, 86, 104001.

Celis-Morales, C., Livingstone, K. M., Marsaux, C. F., Macready, A. L., Fallaize, R., O’Donovan, C. B., Woolhead, C., Forster, H., Walsh, M. C., & Navas-Carretero, S. (2017). Effect of personalized nutrition on health-related behaviour change: evidence from the Food4me European randomized controlled trial. International journal of epidemiology, 46(2), 578–588.

Celis-Morales, C., Marsaux, C. F., Livingstone, K. M., Navas-Carretero, S., San-Cristobal, R., Fallaize, R., Macready, A. L., O’Donovan, C., Woolhead, C., Forster, H., Kolossa, S., Daniel, H., Moschonis, G., Mavrogianni, C., Manios, Y., Surwillo, A., Traczyk, I., Drevon, C. A., Grimaldi, K.,… Mathers, J. C. (2017). Can genetic-based advice help you lose weight? Findings from the Food4Me European randomized controlled trial. Am J Clin Nutr, 105(5), 1204-1213. 10.3945/ajcn.116.145680

Chou, H.-W., Tzeng, W.-C., Chou, Y.-C., Yeh, H.-W., Chang, H.-A., Kao, Y.-C., Huang, S.-Y., Yeh, C.-B., Chiang, W.-S., & Tzeng, N.-S. (2016). Stress, Sleep and Depressive Symptoms in Active Duty Military Personnel. The American Journal of the Medical Sciences, 352(2), 146–153. 10.1016/j.amjms.2016.05.013

de Jong, J. C., Hoevenaars, F. P., Peters, L. G., Berendsen, C. M., Pasman, W. J., Caspers, M. P., Dulos, R., & Wopereis, S. (2026). A Real-Life Digital Intervention for Personalized Nutrition in Adults With Overweight or Obesity: Remote Randomized Controlled Trial. Journal of Medical Internet Research, 28, e73367.

Derossi, A., Husain, A., Caporizzi, R., & Severini, C. (2020). Manufacturing personalized food for people uniqueness. An overview from traditional to emerging technologies. Critical reviews in food science and nutrition, 60(7), 1141–1159. https://www.tandfonline.com/doi/pdf/10.1080/10408398.2018.1559796

Fisher, R. J. (1993). Social desirability bias and the validity of indirect questioning. Journal of consumer research, 20(2), 303–315.

Gevaert, A. B., Adams, V., Bahls, M., Bowen, T. S., Cornelissen, V., Dörr, M., Hansen, D., Kemps, H. M., Leeson, P., & Van Craenenbroeck, E. M. (2020). Towards a personalised approach in exercise-based cardiovascular rehabilitation: How can translational research help? A ‘call to action’from the Section on Secondary Prevention and Cardiac Rehabilitation of the European Association of Preventive Cardiology. European Journal of Preventive Cardiology, 27(13), 1369–1385. https://watermark02.silverchair.com/eurjpc1369.pdf?token=AQECAHi208BE49Ooan9kkhW_Ercy7Dm3ZL_9Cf3qfKAc485ysgAAA4UwggOBBgkqhkiG9w0BBwagggNyMIIDbgIBADCCA2cGCSqGSIb3DQEHATAeBglghkgBZQMEAS4wEQQMQQw2iABN63oXVRfNAgEQgIIDOCUEYh4Ig2V2--buFtrydIPqH2YsNodEQFE6A9PUspsRyYePLVQQUbaiRnvRFS1-NomhVldm2TKRImHIkm8anf7Z8JOleiCGuCHZ5_MFj_GNqAw5vriTPzvVqXsSon5LwR33zWFnlwKoa0rGYyIQFsyD0VTdbEQvb5tEXe4Em1jtz4OIeuaFilQK8C18mv3OSmL-kygU5c3zP4SI2N6pof91LBgECzzL8swSbR_6gFqY36QAhfXgPR9NhOD6js6oZa0nmUVpnr73pVTmayTGkc5jhYaD46tmZZKJS7dQ4dPP08f-Bpw6xLFM-AEBlA1kJXk89R86GTmIXFAZAlKOYjG2JPKJegOoVJvCS8lcgX4E4QocOooQ5JRsWb2fc-7EMDkpfY9rEN8McgtELGlqb9Utb_fTOpkoCATJCSL_4HKqOseB-c8eF8dsXrs7x6BwHytyAMf9IbfcYQG1sUXtuP3rePhmUecqsUZOkfN4-hqZOzeUb8A6wCx313QLBoM319D6G0TPJEcnn05--Gf5zFr6xDFxQi_o9uSOzrwx9rbBQ5RQYHyUASciKi0hR_jYP11FFCLik_YHXnxn-fEmuF9DqL2w6FNdtAtWxfcepD4YVpHZnrfUFwV19yA_oqe7pVGttW0Lt8T8oV7VrBfsN1P1KCD04fF-FnoWKhpx4NgmnIEaycPlEwOFvrI488OcH1nVSb2suSjmzAwfRdUPE5MQlDCwqQbqOQS3iop39V3G04aLrMdfh6R5erpIwPMuAOCgV7NVpzT0LgFQcL2f1_1Qf5NnVVXlbqG1-OIqxYCrxFwK9lYhRDKHUjWzJqpOfMuiQ6Ys4LbvOFBbxGrnMpYTd7z61XrKHWBxLR046pBqK-X2FPEgS0ULxUFJW6NrVXNf3hwUqLCA0tA3czVuJTDUiMgXlA1GWwYaowAzrC2izICPdiQkJc_fyUNSVIdhxbg7unJbp0nIAU6koyfoS3IDmIj-yWFIGKQjAowMYLAsmHBbWlgfT0F9s4LOrtIUty14KELfC-6OtRpXrX8F-MdkGfmfJWHPJwxQ5BE0HajGreCQGX2b8PwuvOdgkQTwXqaFqSsbGjsY

Hauret, K. G., Jones, B. H., Bullock, S. H., Canham-Chervak, M., & Canada, S. (2010). Musculoskeletal injuries: description of an under-recognized injury problem among military personnel. American journal of preventive medicine, 38(1), S61–S70.

Jinnette, R., Narita, A., Manning, B., McNaughton, S. A., Mathers, J. C., & Livingstone, K. M. (2021). Does Personalized Nutrition Advice Improve Dietary Intake in Healthy Adults? A Systematic Review of Randomized Controlled Trials. Advances in Nutrition, 12(3), 657–669. 10.1093/advances/nmaa144

Karl, J. P., Margolis, L. M., Fallowfield, J. L., Child, R. B., Martin, N. M., & McClung, J. P. (2022). Military nutrition research: contemporary issues, state of the science and future directions. European journal of sport science, 22(1), 87–98. https://www.tandfonline.com/doi/10.1080/17461391.2021.1930192?url_ver=Z39.88-2003&rfr_id=ori:rid:crossref.org&rfr_dat=cr_pub 0pubmed

Khemacheevakul, K., Wolodko, J., Nguyen, H., & Wismer, W. (2021). Temporal Sensory Perceptions of Sugar-Reduced 3D Printed Chocolates. Foods, 10(9), 2082. https://www.mdpi.com/2304-8158/10/9/2082

König, L. M., Attig, C., Franke, T., & Renner, B. (2021). Barriers to and facilitators for using nutrition apps: systematic review and conceptual framework. JMIR mHealth and uHealth, 9(6), e20037.

Lyndhurst, B. (2009). An evidence review of public attitudes to emerging food technologies. Social Science Research Unit, Food Standards Agency, Crown, 1, 83.

Michaut, A. (2004). Consumer acceptance of new products with application to foods. Unpublished doctoral dissertation, Wageningen University, Wageningen, the Netherlands.

Podsakoff, P. M., MacKenzie, S. B., Lee, J.-Y., & Podsakoff, N. P. (2003). Common method biases in behavioral research: a critical review of the literature and recommended remedies. Journal of applied psychology, 88(5), 879.

Popa, M. E., & Popa, A. (2012). Consumer behavior: determinants and trends in novel food choice. Novel Technologies in Food Science: Their Impact on Products, Consumer Trends and the Environment, 137–156.

Prochaska, J. O., & DiClemente, C. C. (1982). Transtheoretical therapy: Toward a more integrative model of change. Psychotherapy: theory, research & practice, 19(3), 276.

Reinders, M. J., Bouwman, E. P., Van Den Puttelaar, J., & Verain, M. C. (2020). Consumer acceptance of personalised nutrition: The role of ambivalent feelings and eating context. PLoS One, 15(4), e0231342. https://journals.plos.org/plosone/article/file?id=10.1371/journal.pone.0231342&type=printable

Rijnaarts, I., De Roos, N. M., Wang, T., Zoetendal, E. G., Top, J., Timmer, M., Bouwman, E. P., Hogenelst, K., Witteman, B., & De Wit, N. (2021). Increasing dietary fibre intake in healthy adults using personalised dietary advice compared with general advice: a single-blind randomised controlled trial. Public health nutrition, 24(5), 1117-1128. https://www.cambridge.org/core/services/aop-cambridge-core/content/view/1D6A517830DD040E34BDC60BD49C79EC/S1368980020002980a.pdf/div-class-title-increasing-dietary-fibre-intake-in-healthy-adults-using-personalised-dietary-advice-compared-with-general-advice-a-single-blind-randomised-controlled-trial-div.pdf

Rijnaarts, I., de Roos, N. M., Wang, T., Zoetendal, E. G., Top, J., Timmer, M., Hogenelst, K., Bouwman, E. P., Witteman, B., & de Wit, N. (2022). A high-fibre personalised dietary advice given via a web tool reduces constipation complaints in adults. Journal of Nutritional Science, 11, e31. https://www.cambridge.org/core/services/aop-cambridge-core/content/view/C5E2898A53B8ABCF6B95AE60FE3C7E9F/S2048679022000271a.pdf/div-class-title-a-high-fibre-personalised-dietary-advice-given-via-a-web-tool-reduces-constipation-complaints-in-adults-div.pdf

Ronteltap, A., & van Trijp, H. (2007). Consumer acceptance of personalised nutrition. Genes Nutr, 2(1), 85–87. 10.1007/s12263-007-0003-z

Ross, M. M., Collins, A. M., McCarthy, M. B., & Kelly, A. L. (2022). Overcoming barriers to consumer acceptance of 3D-printed foods in the food service sector. Food Quality and Preference, 100, 104615.

Silva, F., Pereira, T., Mendes, S., Gordo, L., & Gil, M. M. (2024). Consumer’s perceptions and motivations on the consumption of fortified foods and 3D food printing. Future Foods, 10, 100423. 10.1016/j.fufo.2024.100423

Steptoe, A. (1995). Development of a measure of the motives underlying the selection of food: the food choice questionnaire. Appetite.

Sun, J., Zhou, W., Huang, D., Fuh, J. Y., & Hong, G. S. (2015). An overview of 3D printing technologies for food fabrication. Food and bioprocess technology, 8, 1605–1615.

Talens, C., Rios, Y., & Santa Cruz, E. (2022). Leveraging innovative technologies for designing a healthy and personalized breakfast: consumer perception of three smart cooking devices in the EU. Open Research Europe, 1, 151.

Tharion, W. J., Lieberman, H. R., Montain, S. J., Young, A. J., Baker-Fulco, C. J., DeLany, J. P., & Hoyt, R. W. (2005). Energy requirements of military personnel. Appetite, 44(1), 47–65. 10.1016/j.appet.2003.11.010

Thomas, J. L., Wilk, J. E., Riviere, L. A., McGurk, D., Castro, C. A., & Hoge, C. W. (2010). Prevalence of mental health problems and functional impairment among active component and National Guard soldiers 3 and 12 months following combat in Iraq. Archives of general psychiatry, 67(6), 614–623. https://watermark02.silverchair.com/yoa90102_614_623.pdf?token=AQECAHi208BE49Ooan9kkhW_Ercy7Dm3ZL_9Cf3qfKAc485ysgAAAzgwggM0BgkqhkiG9w0BBwagggMlMIIDIQIBADCCAxoGCSqGSIb3DQEHATAeBglghkgBZQMEAS4wEQQMrj8o6-wEc9wG2myRAgEQgIIC63BvPO_xF38-3dyDrDimtKI6JJdIiSi1YHgkIPNcKHXntDHj5yOYr4D0NtEt__vYOFLA4Y0Jbfs2nmxFqz1IhgqPgyPYPs3JjJUXcJFP2x0ONVYyuZiuepUes67KbzaOirAMZd6hRJuajH_In9ez3eJ5uITihVvs9X_D-frdmJT6p9u978JZEMbGjnP0ytI69RVV4VZDicRPB3TpYWSL4vZsfjYr4vR8v0y5l5DKBu7mhMQ3HD9kSkS4DUfBfkZaDDQDt-fav3YV1KoXmuRVWvAssQDmxdk443T8ReQfF2fk61R-ogKsmbzorK2zvbO6tYLNy6lyxxagUVBw4l8HKWr--LjcMjLW1DD9ADTeabDtbQBgw4Qe4TxVUt36rKGb6ZESy5juX5slO-TpYJeDbO_1RZD0Ca7ZNTCqfyMTBJJqVrN-gIl4M16lW6RhPTKPXEAQIxu7zBYs3EOKYVu59D2LZ-PxuVOTwzc96riOOZCD5oKWtI0ADf3kk9RW1CDtOHzAoBLV3HUvUVad4EHfVd2159OA4uZovcJkMf7HwALgGgm8Jb8Fm3ZOz3McCJ-ZaGDQ2GjVJ3aiGZh_lv-c-w8x1lev-E0zMoTE0forJuKzfKJCC6jJUa7dXwDeg_mtMDPlVQSW87fJT2RU6EAikKbtfQMr-te-fIhaGh-dO0zjCGLJ-65wEWUmsvrDeW_GUg-0K8E2kC9FlOsLiVF6rvRxyw4I7Gp4aroJdGaSIBJEFM4oKYfD5HU_KS5zqBabRMVSIlKs9C2Jkeu9P22kxprkiLo7PiEU0HlpBbrqNbcpCGzsfR7ffg--6gvEjUbUwRSidQ8qnfB9K6UlXoNHUzEsRbYXtXfM0af647l582Pvtp9gXgc_nkLBioQlpSjiObuhNSpQEJ1E8X8wlzsL9Rh52SsYJGIpae6lUUd7VHvFo_-F_nM4xF_5smRCDg2cCO56OQmvnbuKoORFJg7e3-PXssMNRByqc-RgIA

Trouwborst, I., Gijbels, A., Jardon, K. M., Siebelink, E., Hul, G. B., Wanders, L., Erdos, B., Péter, S., Singh-Povel, C. M., & de Vogel-van den Bosch, J. (2023). Cardiometabolic health improvements upon dietary intervention are driven by tissue-specific insulin resistance phenotype: a precision nutrition trial. Cell Metabolism, 35(1), 71–83. e75.

van der Haar, S., Hoevenaars, F. P., van den Brink, W. J., van den Broek, T., Timmer, M., Boorsma, A., & Doets, E. L. (2021). Exploring the potential of personalized dietary advice for health improvement in motivated individuals with premetabolic syndrome: pretest-posttest study. JMIR Formative Research, 5(6), e25043.

van der Haar, S., Raaijmakers, I., Verain, M. C., & Meijboom, S. (2023). Incorporating Consumers’ Needs in Nutrition Apps to Promote and Maintain Use: Mixed Methods Study. JMIR mHealth and uHealth, 11(1), e39515.

Wang, W.-F., Guo, X.-X., & Yang, Y.-S. (2015). Gastrointestinal problems in modern wars: clinical features and possible mechanisms. Military Medical Research, 2, 1–8.

Yetley, E. A., MacFarlane, A. J., Greene-Finestone, L. S., Garza, C., Ard, J. D., Atkinson, S. A., Bier, D. M., Carriquiry, A. L., Harlan, W. R., Hattis, D., King, J. C., Krewski, D., O’Connor, D. L., Prentice, R. L., Rodricks, J. V., & Wells, G. A. (2017). Options for basing Dietary Reference Intakes (DRIs) on chronic disease endpoints: report from a joint US-/Canadian-sponsored working group123. The American Journal of Clinical Nutrition, 105(1), 249S–285S. 10.3945/ajcn.116.139097

Zeevi, D., Korem, T., Zmora, N., Israeli, D., Rothschild, D., Weinberger, A., Ben-Yacov, O., Lador, D., Avnit-Sagi, T., & Lotan-Pompan, M. (2015). Personalized nutrition by prediction of glycemic responses. Cell, 163(5), 1079–1094.

Zhang, L., Noort, M., & van Bommel, K. (2022). Towards the creation of personalized bakery products using 3D food printing. In Advances in food and nutrition research (Vol. 99, pp. 1–35). Elsevier.

