## Supplemental tables 1a1b for "Implementing a Personalised Nutrition Service in a Real-Life Military Setting: Feasibility and Consumer Acceptance"

**Supplemental Table 1a: Nutritional composition of the Personalised Nutrition Product (PNP) health benefits**

| **Health benefit** | **Dough** | **Filling** | **Reasoning for Dough** **– Filling combination** | **Micronutrients** | **Reasoning for micronutrients** |
| --- | --- | --- | --- | --- | --- |
| **Endurance** | Cellular ink (Carrot) | Ganache | Complex carbohydrates and high energy content to support endurance exercise | Caffeine | Sports nutrition position stand on caffeine use for endurance (5,6,9) |
| **Muscle Power** | Cereal | Yoghurt | Carbohydrates to support energy demands of the activity | Beta alanine | Can increase power output and capacity (10, 11) |
| **Recovery** | Cereal, almond | Custard | Sports nutrition Position stand on protein (12) | Leucine | Sports nutrition Position stand on Leucine (12) |
| **Immune Health – Endomorph bodytype** | Cereal, high fibre | Banana-strawberry | Fibers to support immune health (3), combination supports endomorph macronutrient profile | Zinc, Vitamin C, Folic acid | EFSA health claims on Zinc, Vit C and Folic acid to support the immune system (7,8, 14) |
| **Immune Health -Mesomorph and Ectomorph bodytype** | Cereal, almond | Apple | Fibers (3) and Poly unsaturated fatty acids (4) to support immune health, combination supports mesomorph and ectomorph macronutrient profile | Zinc, Vitamin C, Folic acid | EFSA health claims on Zinc, Vit C and Folic acid to support the immune system (7,8,14) |
| **Mental Performance – Endomorph bodytype** | Protein | Peanut butter | combination supports endomorph macronutrient profile | Caffeine, Folic acid | Sports nutrition position stand on caffeine use for cognitive function including attention and vigilance (5), EFSA health claims on folic acid to reduce tiredness & fatigue and to support normal psychological function |
| **Mental Performance – Mesomorph bodytype** | Cellular ink (plum – no added sugar) | Peanut butter | combination supports mesomorph macronutrient profile | Caffeine, Folic acid | Sports nutrition position stand on caffeine use for cognitive function including attention and vigilance (5), EFSA health claims on folic acid to reduce tiredness & fatigue and to support normal psychological function |
| **Mental Performance – Ectomorph bodytype** | Protein | Custard | combination supports ectomorph macronutrient profile | Caffeine, Folic acid | Sports nutrition position stand on caffeine use for cognitive function including attention and vigilance (5), EFSA health claims on folic acid to reduce tiredness & fatigue and to support normal psychological function |
| **Digestive Health – Endomorph bodytype** | Cereal, high fibre | Peanut butter | Fibers to support digestive health (18), combination supports endomorph macronutrient profile | Probiotics | Probiotics support intestinal health (15, 16,17) |
| **Digestive Health – Mesomorph bodytype** | Cellular ink (plum – no added sugar) | Custard | Fibers to support digestive health (18), combination supports mesomorph macronutrient profile | Probiotics | Probiotics support intestinal health (15, 16,17) |
| **Digestive Health – Ectomorph bodytype** | Cellular ink (plum – no added sugar) | Ganache | Fibers to support digestive health (18), combination supports ectomorph macronutrient profile | Probiotics | Probiotics support intestinal health (15, 16,17) |

**Supplemental Table 1b: Composition of the PNPs per health benefit and filling**

| **Health benefit** | **Dough** | **Estimated Weight, g** | **Filling options** | **Energy, Kcal/100g** | **Carbohydrates**  **g/100g** | **Protein, g/100g** | **Fat g/100g** | **PUFA g/100g** | **Fiber, g/100g** | **Micronutriënt enrichment (mg/pp)** |
| --- | --- | --- | --- | --- | --- | --- | --- | --- | --- | --- |
| **Endurance** | Cellular Ink (Carrot) | 100 g | Ganache | 365 | 50 | 3,3 | 14 | 3,53 | 5,6 | Caffeine (192) |
| **Muscle Power** | Cereal | 100 g | Yoghurt | 369 | 53 | 5,9 | 14 | - | 4,3 | Beta alanine (3094) |
| **Recovery** | Cereal, almond | 100 g | Custard | 351 | 49 | 7,9^1^ | 13 | - | 5,1 | Leucine (2200) |
| **Immune Health** |  |  |  |  |  |  |  |  |  |  |
| **– Endomorph bodytype** | Cereal, high fibre | 40 - 70 g | Banana-strawberry | 97 | 14 | 6,5 | 4,4 | 0,0025 | 3,4 | Folic acid (48) , Zinc (52), Vitamin C (196) |
| **-Mesomorph bodytype** | Cereal, almond | 70 g | Apple | 317 | 39 | 1,6 | 8,8 |  | 13 | Folic acid (48) , Zinc (52), Vitamin C (196) |
| **-Ectomorph bodytype** | Cereal, almond | 100 g |  | 169 | 49 | 3,1 | 13 | 0,064 | 5,4 | Folic acid (48) , Zinc (52), Vitamin C (196) |
| **Mental Performance** |  |  |  |  |  |  |  |  |  |  |
| **– Endomorph bodytype** | Protein | 70 g | Peanut butter | 188 | 9,4 | 13 | 12 | 3,42 | 1,3 | Caffeine (192), Folic acid (48) |
| **– Mesomorph bodytype** | Cellular ink (plum – no added sugar) | 70-100 g | Peanut butter | 283 | 28 | 8,1 | 16 | 4,26 | 5,6 | Caffeine (192), Folic acid (48) |
| **– Ectomorph bodytype** | Protein | 100 g | Custard | 281 | 38 | 25 | 3,6 | 1,86 | 2,9 | Caffeine (192), Folic acid (48) |
| **Digestive Health** |  |  |  |  |  |  |  |  |  |  |
| **– Endomorph bodytype** | Cereal, high fibre | 70 g | Peanut butter | 97 | 14 | 1,4 | 15 | 3,42 | 2,3 | Probiotics: Winclove Probiotics (NL) (3094) |
| **– Mesomorph bodytype** | Cellular ink (plum – no added sugar) | 70 – 100 g | Custard | 243 | 35 | 5,4 | 0,33 | 0,064 | 3,7 | Probiotics: Winclove Probiotics (NL) (3094) |
| **– Ectomorph bodytype** | Cellular ink (plum – no added sugar) | 100 g | Ganache | 318 | 48 | 3,8 | 14 | 3,62 | 17 | Probiotics: Winclove Probiotics (NL) (3094) |

^1^ a casing naturally contains 2169mg Leucine per product

3. Venter C, Meyer RW, Greenhawt M, Pali-Schöll I, Nwaru B, Roduit C, et al. Role of dietary fiber in promoting immune health-An EAACI position paper. Allergy. 2022;77(11):3185-98.

4. Jäger, R., Heileson, J. L., Abou Sawan, S., Dickerson, B. L., Leonard, M., Kreider, R. B., … Antonio, J. (2025). International Society of Sports Nutrition Position Stand: Long-Chain Omega-3 Polyunsaturated Fatty Acids. Journal of the International Society of Sports Nutrition, 22(1). <https://doi.org/10.1080/15502783.2024.2441775>

5. Guest NS, VanDusseldorp TA, Nelson MT, Grgic J, Schoenfeld BJ, Jenkins NDM, Arent SM, Antonio J, Stout JR, Trexler ET, Smith-Ryan AE, Goldstein ER, Kalman DS, Campbell BI. International society of sports nutrition position stand: caffeine and exercise performance. J Int Soc Sports Nutr. 2021 Jan 2;18(1):1. doi: 10.1186/s12970-020-00383-4. PMID: 33388079; PMCID: PMC7777221.

6. Spriet LL. Exercise and sport performance with low doses of caffeine. Sports Med. 2014 Nov;44 Suppl 2(Suppl 2):S175-84. doi: 10.1007/s40279-014-0257-8. PMID: 25355191; PMCID: PMC4213371.

9. Pickering, C., Kiely, J. Are the Current Guidelines on Caffeine Use in Sport Optimal for Everyone? Inter-individual Variation in Caffeine Ergogenicity, and a Move Towards Personalised Sports Nutrition. Sports Med 48, 7–16 (2018).

10. Culbertson JY, Kreider RB, Greenwood M, Cooke M. Effects of beta-alanine on muscle carnosine and exercise performance: a review of the current literature. Nutrients. 2010 Jan;2(1):75-98. doi: 10.3390/nu2010075. Epub 2010 Jan 25. PMID: 22253993; PMCID: PMC3257613.

11. Hoffman JR, Stout JR, Harris RC, Moran DS. β-Alanine supplementation and military performance. Amino Acids. 2015 Dec;47(12):2463-74. doi: 10.1007/s00726-015-2051-9. Epub 2015 Jul 24. PMID: 26206727; PMCID: PMC4633445.

12. Jäger, R., Kerksick, C. M., Campbell, B. I., Cribb, P. J., Wells, S. D., Skwiat, T. M., … Antonio, J. (2017). International Society of Sports Nutrition Position Stand: protein and exercise. Journal of the International Society of Sports Nutrition, 14(1). <https://doi.org/10.1186/s12970-017-0177-8>

13. Gonzalez DE, McAllister MJ, Waldman HS, Ferrando AA, Joyce J, Barringer ND, Dawes JJ, Kieffer AJ, Harvey T, Kerksick CM, Stout JR, Ziegenfuss TN, Zapp A, Tartar JL, Heileson JL, VanDusseldorp TA, Kalman DS, Campbell BI, Antonio J, Kreider RB. International society of sports nutrition position stand: tactical athlete nutrition. J Int Soc Sports Nutr. 2022 Jun 23;19(1):267-315. doi: 10.1080/15502783.2022.2086017. PMID: 35813846; PMCID: PMC9261739.

15. Miller LE, Zimmermann AK, Ouwehand AC. Contemporary meta-analysis of short-term probiotic consumption on gastrointestinal transit. World J Gastroenterol 2016; 22(21): 5122-5131

16. Goodman, C., Keating, G., Georgousopoulou, E., Hespe, C., & Levett, K. (2021). Probiotics for the prevention of antibiotic-associated diarrhoea: a systematic review and meta-analysis. BMJ Open, 11. <https://doi.org/10.1136/bmjopen-2020-043054>.

17. Zheng, Y., Zhang, Z., Tang, P., Wu, Y., Zhang, A., Li, D., Wang, C., Wan, J., Yao, H., & Yuan, C. (2023). Probiotics fortify intestinal barrier function: a systematic review and meta-analysis of randomized trials. Frontiers in Immunology, 14. <https://doi.org/10.3389/fimmu.2023.1143548>.

18. Hughes, R., & Holscher, H. (2021). Fueling Gut Microbes: A Review of the Interaction between Diet, Exercise, and the Gut Microbiota in Athletes. Advances in Nutrition, 12, 2190 - 2215. https://doi.org/10.1093/advances/nmab077.
